# Mass drug administration coverage and determinants of drug uptake for elimination of Onchocerciasis in Ulanga District

**DOI:** 10.1101/2024.05.16.24307467

**Authors:** Ambakisye Mhiche, Dinah Gasarasi, George Kabona, Ally Hussein, Ahmed Mohamed Abade

## Abstract

**Background:** Onchocerciasis remain to be an important public health problem targeted for elimination in Tanzania. Ulanga District was known for its high endemicity since 1960s and has been implementing MDA through Community Directed Treatment with Ivermectin (CDTI) strategy since 1998. However, current reports indicate high prevalence of Onchocerciasis in both human and vector species probably because of poor treatment coverage indicating limited evidence for transmission. This study was conducted to assess treatment coverage and explore determinants of drug uptake during MDA program.

**Methods:** A cross-sectional community-based study using multistage cluster sampling was carried out in Ulanga District, Morogoro Tanzania from April-June 2019. Study participants were randomly selected from households and interviewed using a structured questionnaire. Modified Poisson regression was performed to determine independent factors associated with MDA uptake.

**Results:** A total of 502 participants were recruited during the study period with a response rate of 96%. The majority (67%) were females, and the mean age of the study participants was 37.8 ±15 years with an age range of 25–34 (25.5 %). MDA coverage for the studied villages was as follows; Mawasiliano (68%) Uponera (83%), Isongo (84%) and Togo (79%). The drug uptake for all villages was below the optimal coverage recommended by WHO (85%) for successful transmission interruption. Having an age of ≤ 24 years [Adjusted Prevalence Ration (APR) = 3.9(95% CI:1.9-8.3), p < 0.05)], Living in the village for at least a year [APR = 3.4 (95% CI:2.4-4.8), p <0.05)] and believing IVM prevent Onchocerciasis [APR = 13.4(95% CI:2.9-60.9)], p<0.05) were associated with increased chances of Ivermectin uptake during MDA. In addition, decreased drug uptake [APR = 12(95% CI: 2.4-60.9), p<0.05)] was attributable to fear of restriction to drinking alcohol after taking drugs.

**Conclusion:** Low coverage below the WHO optimal recommended coverage has been demonstrated in the studied villages. This implies low drug uptake, delayed interruption of transmission and Onchocerciasis elimination. These findings, therefore, emphasize the need to intensify the MDA awareness campaigns targeting less compliant groups in the community to reinforce the benefits of ivermectin in Onchocerciasis control and address the community misconceptions about MDA.

## Introduction

Onchocerciasis (river blindness) is a vector-borne parasitic disease caused by, *Onchocerca volvulus*, and transmitted by the bites of black flies belonging to *Simulium species* (1). The flies breed in fast-flowing waters of streams and rivers, most notably in Africa, and in Tanzania in particular (2). Onchocerciasis Control focuses on Community Directed Treatment with Ivermectin (CDTI) strategy, consisting of yearly mass drug administration of Ivermectin. Improvement in the ivermectin treatment coverage during MDA have been linked to successful transmission interruption and prevalence reduction in several Onchocerciasis endemic areas (3–5). Onchocerciasis elimination requires 100% geographical coverage in all areas in which transmission occurs, attain and maintain the recommended MDA treatment coverage and demonstrate the interruption of transmission among vector species (6).

Although treatment coverage is an important factor for onchocerciasis elimination process, it only highlights the prevailing situation at community level and does not necessarily guarantee good adherence to drug uptake by individuals in the community. Studies have shown that, individuals who don’t take ivermectin each year provide a sources of reinfection for their communities (7). Therefore, for successful control and elimination of Onchocerciasis all residents in the high-risk population even those who are apparently in good health must take the drugs during the MDA program (6,8–10).

In Tanzania, Onchocerciasis continues to be an important public health problem and it is among the five most prevalent Neglected Tropical Diseases (NTDs) targeted for elimination in the country (11,12). Substantial progress in the control of the diseases has been made and a goal to eliminate Onchocerciasis by 2025 has been set (13). However, to achieve this, community adherence to drug uptake during MDA campaign is of paramount importance.

Ulanga District within Mahenge Onchocerciasis foci in Tanzania was known for its high endemicity since 1960s and has been implementing MDA through CDTI strategy since 1998(14). Despite two decades of MDA in the area transmission is still ongoing raising uncertainty on its possibility to control and eventually eliminate the disease (15). According 2009 and 2017 survey data, Ulanga had an overall maximum prevalence of 22%, Onchocerciasis nodule prevalence was 2.3%, while the positivity rate for OV16 rapid test was 76.5% and the risk of transmission among children aged 6-9 years was 20.7%. Existence of co-endemic conditions such as Epilepsy implies presence of relatively high level of filaria worms in the area, suggesting limited progress made in the control of Onchocerciasis (17). On the other side, literature shows the black fly carries infective parasites and the infection rate is clearly above the threshold for transmission interruption within Ulanga district (18). Therefore, this study was designed to assess the coverage and determinants for drug uptake for Ivermectin MDA in Ulanga district in order to gather appropriate evidence to inform all stakeholders involved in the endeavor to fight Onchocerciasis in the country.

## Methods

### Study Area

The study was conducted in Morogoro region within Mahenge Onchocerciasis focus at Ulanga district council. The area is in southeastern Tanzania; it is a mountainous area with fast-flowing perennial rivers which are Luli, Mbalu, Lukande, Mzelezi, Ruaha and Msingizi Rivers that serves as the potential breeding sites for black flies. The 2012 population census reported 265,203 people living in Ulanga district. Administratively the district has been divided into 3 divisions and 31 wards whereby each ward has about three and above villages. The district OV16 prevalence stands at 76% and 50% in rural and suburban areas respectively. Evidence of active transmission among children aged 6-9 years has been recorded to be 20.7% (17). (**Figure 1**)

### Study Design

This was a cross-sectional community-based study using a multistage cluster sampling method involving both secondary data review and survey carried out in Ulanga district, Morogoro region from April-June 2019.

### Study Population

The study was done on community members with ≥15 years of age who were found at household of the sampled villages. Individuals who had participated in the MDA program for at least one (1) year were eligible if were able explain themselves clearly. The study excluded Individuals who were exempted during MDA campaign because of ineligibility such as being below 5 years old, pregnant women, and those with serious health problems.

### Sample size Estimate

Multistage cluster sampling method was used, while the number clusters and design effects for cluster sampling was determined using Steve Bennett’s, the sample size was determined using the Kish-Leslie formula for single proportion cross sectional study. Sample size calculation considered the assumption that proportion of drug uptake during MDA in Mahenge would be 78% as previously determined. With a 5% level of precision, standard critical value of 1.96 with 95% confidence interval, and non-response rate of 10%.

Design effects and number of clusters were determined using Steve Bennett’s formula as shown below(69).

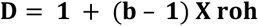

Were; D= Design effect, b= expected number of households covered in each sub-village (20) and roh = Rate of homogeneity among clusters (0.1)

Then, D= 1+ (20 - 1) x 0.1 hence D= 2.9

Number of clusters (Hamlet/Sub-village) was calculated using Steve Bennett’s formula (69)given below

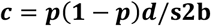

Where, C= Number of clusters (hamlet/sub-villages) that will be included in the study, p = Proportion drug uptake in Ulanga (78%), D=Design effect (2.9), s=Level of precision (5%) and b=expected number of households covered in each sub-village (35)

Then, C = 0. 78 x (1-0.78) x 2.9 and 0.052 × 35 hence C = 5.7 ∼ 6

Determination of sample size was done based Kish-Leslie Formula for single proportion cross sectional study as indicated below.

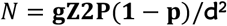

Where: N = Sample size, Z= standard critical value, P= Proportion drug uptake in Ulanga and d=The acceptable margin of error to be 5%

Then; N = [2.9 × 1.962 × 0.78 (1-0.78)] ÷ 0.0652, N = [2.9 × 1.962 × 0.78x 0.22] ÷0.0652 hence N = 453

After adjusting for non -response rate of 10%, 503 was obtained as the minimum sample size.

### Data Collection Procedures

The study involved primary data collection households randomly selected from 4 cluster sampled villages. First stage: Ulanga District council was selected as one of the councils in Mahenge Onchocerciasis foci with high active transmission of Onchocerciasis despite ongoing MDA intervention. Second stage: a list of onchocerciasis endemic wards in Mahenge division was obtained from DMO office, two wards were randomly selected. Third stage: a list of the villages from the selected wards were obtained and four villages were randomly selected. From which, 6 hamlets were selected according to Steve Bennett’s formula. Fourth stages: households from each selected hamlet were randomly selected with starting point at the centre of the hamlet; each eligible household member was prospectively followed until the anticipated sample size of 503 was attained.

Secondary data collection was done in 2 health facilities within the selected villages. A tool to assess the MDA uptake was developed to collect information among those who consented to participate. Apart from questionnaire, a checklist for data abstraction from community drug distributers register (community register) was also developed. In addition, an interview guide was developed to guide the key informant interviews. An Open Data Kit (ODK Collect) mobile application freely available in google play (https://play.google.com/store/apps/details?id=org.odk.collect.android&hl=en&gl=US) was developed and used to collect data. The data collection tools were pre-tested to assess the adequacy of the interviewer-based questionnaire and the accuracy of mastering data entry using a mobile phone devise as well as to ascertain their validity and reliability.

The necessary permissions to conduct the study were sought from respective authorities (region, district, ward and village governments). Data were collected using mobile devices through Open Data Kit (ODK Collect) application. Questionnaire were uploaded into mobile phone, corrected and checked for quality within the respective mobile devises daily by the Information Technology (IT) personnel from The National NTDs Control Program. Review of all Community Drug Distributors register in the selected villages was done to determine individuals in the community who participated in the past MDA campaigns using a checklist. Interviewer administered semi structured questionnaire was used to collect information on factors that determine drug uptake in the community. The questionnaire had three (3) major parts; part A had 7 questions that explored the socio-demographic characteristics of the participants. Part B had 13 questions that explored the community perceived disease and drug effects that influence drug uptake especially treatment effects, perceived knowledge, belief and perception about Ivermectin drugs. Part C had 3 questions that looked at the program expert support, awareness creation and drug supply system for ivermectin distribution particularly community sensitization both formal and informal in facilitating individual decision on taking of ivermectin drugs during MDA campaign. Data completeness and accuracy was checked on daily basis and any ambiguities were immediately addressed the following day during the study period April to June 2019.

### Data Management and Analysis

Data were transferred to the NTDCP server from the mobile phone (Smartphone) after crosschecking their completeness and accuracy. From the server data were extracted as excel files that could be easily read by Epi Info and STATAanalysis software. Data was cleaned and analyzed using both Epi Info 7.2.2.6 by CDC Atlanta, Georgia (US) and Stata 15 by College Station, Tx:StataCorpLLC. Confidence Intervals around proportions were calculated using Fleiss Quadratic approximation formula and are reported at 95% level. Following descriptive analysis, generalized linear model with modified Poisson regression was used to estimate prevalence ratio. The variables were further subjected to Poisson regression model with negative binomial link robust. The variables with p≤0.05 were regarded as determinants of drug uptake. MDA drug Uptake, Socio demographic, drug consumer behavior, CDD behavior and IVM treatment factors were analyzed and summarized in figure 1 and Tables 1 and 2. Community and program delivery factors were categorized based on Key informants coded information.

**Table 1:**
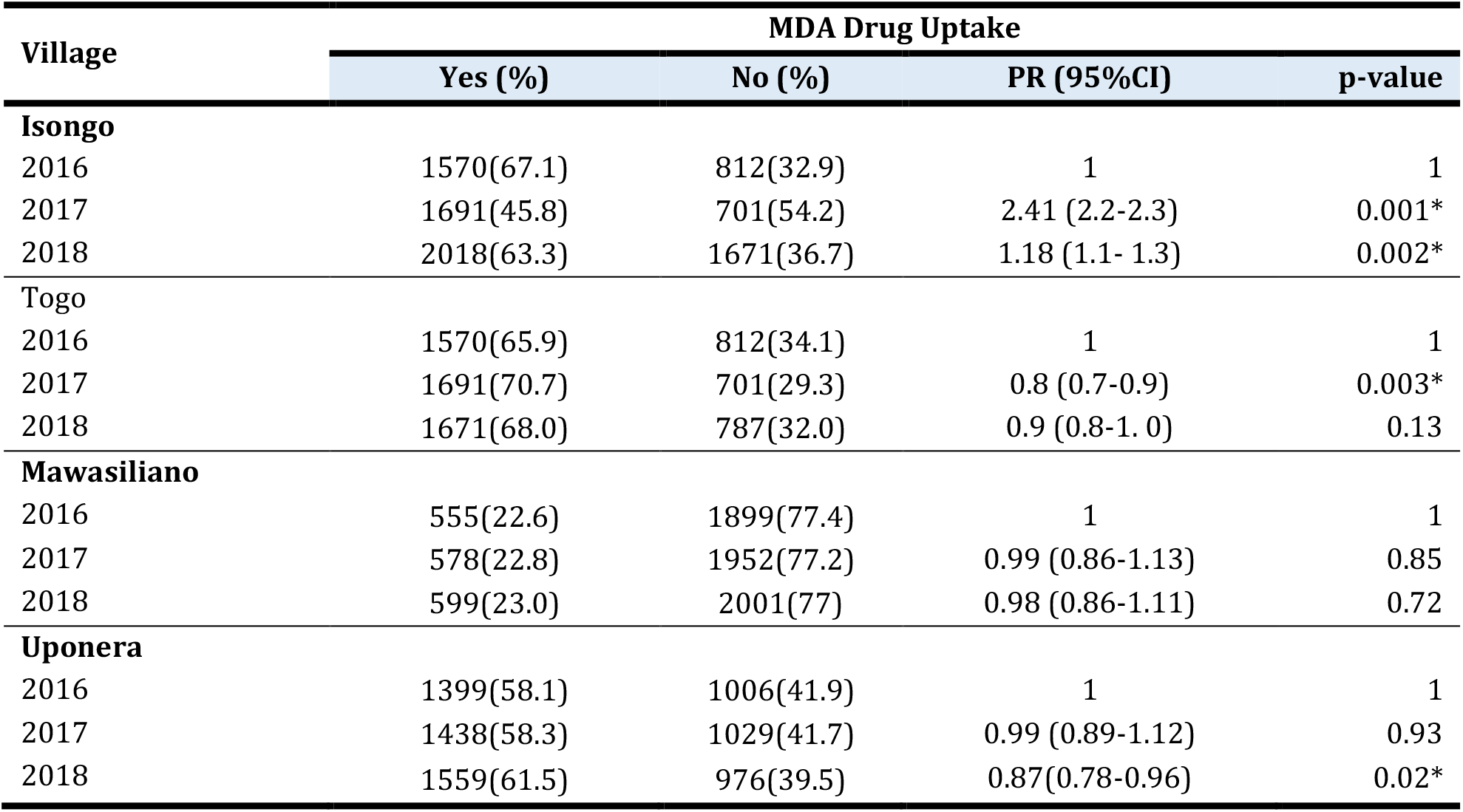
Comparison of Drug uptake along three-year period in selected villages in Ulanga District.

**Table 2:** Socio demographic characteristic associated with MDA drug uptake in multi variate analysis, in Ulanga district, June 2019.

| Variable | MDA Drug Uptake |  |  |  |  |  |
| --- | --- | --- | --- | --- | --- | --- |
|  | Yes (%) | No (%) | CPR (95%CI) | p-value | APR (95%CI) | p-value |
| **Age:** |  |  |  |  |  |  |
| 15-24 | 72 (64.3) | 40(35.7) | 3.9(1.9-8.3) | 0.00* | 2.8(1.3-6.2) | 0.008* |
| 25-34 | 94 (76.4) | 29 (23.6) | 2.6(1.2-5.6) | 0.02* | 2.3(1.04-4.9) | 0.04* |
| 35-44 | 80(82.50) | 17(17.5) | 1.9(0.8-4.4) | 0.12 | 1.9(0.8-4.3) | 0.13 |
| 55 - 93 | 64(87.7) | 9(12.3) | 1.4(0.5- 3.5) | 0.52 | 1.3(0.5-3.4) | 0.53 |
| 45-54 | 70(90.9) | 7(9.1) | 1 | 1 | 1 | 1 |
| **Education Level:** |  |  |  |  |  |  |
| None | 16 (69.6)) | 7 (33.4) | 1.4(0.7- 3.9) | 0.25 | 1.7(0.6-4.6) | 0.29 |
| Secondary | 59 (67.1) | 29 (32.9) | 1.1(0.7-1.7) | 0.72 | 1.1(.6-2.2) | 0.68 |
| College | 8(66.7) | 4(33.3) | 1.7(0.7-8.2) | 0.16 | 2.1(0.4-10.4) | 0.37 |
| Primary | 297(82.7) | 62(27.3) | 1 | 1 | 1 | 1 |
| **Marital Status** |  |  |  |  |  |  |
| Married/Cohabiting | 230(83) | 47(17) | 1 | 1 | 1 | 1 |
| Not Married | 150(73.2) | 55(26.8) | 1.2(1.2-2.8) | 0.009* | 1.2(0.8-1.7) | 0.39 |
| **Occupation** |  |  |  |  |  |  |
| Peasant | 341 (81.2) | 79 (18.8) | 1.9 (0.4-8.2) | 0.42 | 1.8(0.6-5.4) | 0.30 |
| SMEs | 14(58.3) | 10(41.7) | 5.7(1.1-30.6) | 0.04* | 3.2(0.99-10.6) | 0.05 |
| Student | 9(45) | 11(55) | 9.8(1.8-54.3) | 0.009* | 2.9(0.9-9.8) | 0.09 |
| Extension Worker | 16(88.9) | 2(11.1) | 1 | 1 | 1 | 1 |
| **Duration of stay in the village** |  |  |  |  |  |  |
| Below one year | 10(37) | 17(63) | 1 | 1 | 1 | 1 |
| One year and above | 370(81.3) | 85(18.7) | 7.4(3.3-16.7) | 0.0001* | 2.3(1.5-3.4) | 0.00* |

### Ethical consideration

Ethical clearance with reference.No.DA.287/298/01A was obtained from the Muhimbili University of Health and Allied Sciences, Research and Publications Ethical Committee. Research permits were obtained from Regional Administrative Secretary for Morogoro region and later by the District Executive Director for Ulanga District council. Confidentiality was secured during the data collection, thus name and addresses of the patient were not recorded in the mobile data collection application. Only the Programmer IT and Principle Investigator had password to access data on the server. Written informed consent was obtained from willing respondents prior to data collection. Semi structured and in-depth Interviews were conducted strictly in a private place. Individuals not willing to participate after informed consent were excluded and this did not affect in any way their participation in future MDA campaign.

## Results

A total of 502 participants were recruited during the study period with a response rate of 96%. The mean age of the study participants was 37.8 ±15 years majority being in the age range of 25–34 (25.5 %) while Females were 67%. MDA coverage was 68%, 83%, 84% and 79% for Mawasiliano, Uponera, Isongo and Togo villages respectively (Figure 1). Drug uptake coverage from all studied villages were below the optimal coverage (85%) recommended by WHO for successful transmission interruption and elimination of Onchocerciasis. (Figure 1). Drug uptake coverage observed in Isongo, Togo and Uponera was significantly different for the three-year period. While there was no significant difference in drug uptake along three-year period for Mawasiliano villages. In Isongo village, drug uptake was 2.4 times higher in 2017 [PR = 2.41 (2.2-2.3)], While drug uptake in 2018 was 1.2 times higher, [PR = 1.18 (1.1-1.3)] compared to that of 2016 respectively. On the other hand, In Uponera Village compliance to drug uptake was 0.9 times less in 2018 compared to 2016, [OR = 0.87(0.78-0.96)], **(Table 1)**

### Socio demographic Factors associated with MDA drug uptake

A stepwise modified Poisson regression model was developed that included Age, Education level, Marital Status, Occupation and Duration of stay in the villages to determine their association with drug uptake. Age and duration of stay in the village among participants were found to be independent predictors of drug uptake. Participants aged 15-24 and 25-34 years was 3 times [APR = 2.8(95% CI:1.3-6.2)] and 2 times [APR = 2.3(95% CI:1.04-4.9)] higher respectively, compared to adult population ages 45-54. (Table 2). Drug uptake among participants who had lived in the district for at least a year was 2 times higher compared to those who had lived in the district for less than a year. [APR = 2.3(95% CI: 1.5-3.4)] **(Table 2)**.

### Perceived Disease and Drug Effects and their Influence on Drug Uptake

Several factors such as pre-MDA sensitization, MDA distribution cycle, perceived benefit of IVM in symptoms control, understanding MDA distribution interval and perceived reason for drug uptake among others) were investigated as to their association with drug uptake among communities in Ulanga. Drug uptake among participants who didn’t know about the MDA distribution cycle was found to be more than two times less compared to participants who knew the distribution cycle was once a year in the district [APR = 4.72.5(95% CI: 1.1-6.0)]. Those who believed that Ivermectin prevents Onchocerciasis were 13 times more likely to take the drug compared to those who feared Ivermectin side effects [APR = 13.4(95% CI:2.9-60.9)]. Drug uptake among participants who refrained from Ivermectin because of alcohol drinking behavior was found to be 12 times less compared to those whose drug uptake was influenced by ivermectin side effects [APR = 12(95% CI: 2.4-60.9)] **(Table 3)**.

**Table 3:** Factors associated with MDA drug uptake in multi variate analysis, in Ulanga district.

| Variable | MDA Drug Uptake |  |  |  |  |  |
| --- | --- | --- | --- | --- | --- | --- |
|  | Yes | NO | CPR (95%CI) | P-value | APR (95%CI) | P-value |
| **Understanding Onchocerciasis Symptoms** |  |  |  |  |  |  |
| Knows symptoms | 357(86.6) | 55(13.4) | 5.0(3.7-6.8) | 0.00* | 0.9(0.8-1.1) | 0.38 |
| Don't know the symptoms | 23(76.7) | 47(23.3) | 1 | 1 | 1 | 1 |
| **Pre MDA advocacy and sensitization attended** |  |  |  |  |  |  |
| No | 208(92.9) | 16(7.1) | 4.7(2.8-7.1) | 0.00* | 1.02(0.8-1.3) | 0.85 |
| Yes | 172 (66.3) | 86(33.3) | 1 | 1 | 1 | 1 |
| **Onchocerciasis Targeted health education** |  |  |  |  |  |  |
| Hygiene and Ocho | 118(98/3) | 2(1.7) | 0.2(0.05-1.13) | 0.07 | 0.4(0.2-1.2) | 0.12 |
| Other but not Ocho | 78(57.1) | 6(42.9) | 6(2.2-16.0) | 0.01* | 0.6(0.2-1.7) | 0.33 |
| Don't remember | 176(66.7) | 88(33.3) | 4.7((2.1-10.3) | 0.00* | 0.6(0.2-1.8) | 0.36 |
| Onchocerciasis only | 78(92.9) | 6(7.1) | 1 | 1 | 1 | 1 |
| **Perceived impact of advocacy attended in MDA drug uptake.** |  |  |  |  |  |  |
| Didn't Influenced | 195(97.5) | 5(2.5) | 1 | 1 | 1 | 1 |
| Influenced next MDA | 185(65.6) | 97(34.4) | 13.8(5.7-33.2) | 0.00* | 2.1(0.7-6.4) | 0.17 |
| **Source Pre uptake Health Education** |  |  |  |  |  |  |
| FLHW | 184(94.4) | 11(5.6) | 1.4(0.3-6.2) | 0.65 | 1.5(0.8-2.6) | 0.18 |
| TV and Radio | 52(80) | 13(20) | 5(1.2- 21.2) | 0.03* | 1.6(0.9-2.5) | 0.07 |
| Never heard | 96(55.8) | 76(44.2) | 11(2.8-43.5) | 0.00* | 1.6(0.9-2.6) | 0.05 |
|  | Yes | NO | CPR (95%CI) | P-value | APR (95%CI) | P-value |
| Reference CDD | 48(96) | 2(4) | 1 | 1 | 1 | 1 |
| **Benefit of IVM in symptoms control** |  |  |  |  |  |  |
| Stop Itching | 325(97.3) | 9(2.7) | 1 | 1 | 1 | 1 |
| Don't Stop Itching | 37(97.4) | 1(2.6) | 0.97(0.12-7.5) | 0.98 | 2.6(0.3-20.4) | 0.38 |
| Don't know | 18 (16.4) | 92(83.6) | 31(16.2-59.5) | 0.00* | 4.7(1.03-21.1) | 0.05* |
| **Perceived fear of IVM side effects** |  |  |  |  |  |  |
| Influenced drug uptake | 83(67.5) | 40(32.5) | 1.8(1.4-2.6) | 0.00* | 1.2(0.9-1.3) | 0.08 |
| Not influenced drug uptake | 297(82.7) | 40(17.3) | 1 | 1 | 1 | 1 |
| **Experience of previously treated patient** |  |  |  |  |  |  |
| Influenced drug uptake | 244(75.8) | 78(24.2) | 1.6(1.1-2.5) | 0.02* | 1.1(0.9-1.3) | 0.15 |
| Not influenced drug uptake | 136(85) | 24(15) | 1 | 1 | 1 | 1 |
| **Understanding MDA distribution Interval** |  |  |  |  |  |  |
| Once a year | 311(96.6) | 11 (3.4) | 1 | 1 | 1 | 1 |
| After every 6 months | 62(89.9) | 7(10.1) | 2.9(1.2- 7.4) | 0.02* | 1.3(0.3-5.5) | 0.73 |
| Don't Know | 7(7.7) | 82(92.3) | 27 (15.1-48.5) | 0.00* | 2.5(1.1-6.0) | 0.03* |
| **History of previous effects following drug uptake** |  |  |  |  |  |  |
| At least two | 267(88.1) | 36(18.9) | 1 | 1 | 1 | 1 |
| One Effect | 113(63.1) | 66(36.9) | 3.1(2.1- 4.5) | 0.00* | 0.9(0.8-1.1) | 0.56 |
| **Perceived reason for drug uptake:** |  |  |  |  |  |  |
| Fear of alcohol restriction | 81(92) | 7(8) | 4.8(1.5-14.6) | 0.007* | 12(2.4-60.9) | 0.003* |
| Prevention Effects | 5(5.3) | 90(94.7) | 56.7(23.7-135.4) | 0.00* | 13.4 (2.9-60.9) | 0.001* |
| Side effects | 294(98.3) | 5(1.7) | 1 | 1 | 1 | 1 |

## Discussion

Although MDA has been implemented in Ulanga district for the past 20 years there is still low compliance to drug uptake by individuals in the community. There are still uncertainties over the reported treatment coverage and the relative contribution of various programmatic and social demographic factors in sustaining optimal coverage leading to transmission interruption. This study focused on assessing reported treatment coverage and determining the factors associated with drug uptake in the community as potential explanatory factors for While participants aged 15-24 years had increased chances of drug uptake there has been no significant changes in drug uptake among people aged 35-44 and ≥ 55 in comparison to those aged 45-54 (Table 2). This could due to the fact that people who are aged 15-24 years are either at primary or secondary school level of education, therefore easily accessible through the school system(41,64). There has been increased proportion of people who do not participate in MDA drug uptake as you move across different age group, suggesting potential increase in the non-compliance to drug uptake in the community unlike findings from other studies that reported no significant difference in drug uptake across age groups (64,65). Low drug uptake among participants who had lived in the area for less than one year could be due to inadequate knowledge about Onchocerciasis, which may be attributable to minimal risk perception about the diseases and low awareness about MDA campaign. This finding is similar to other studies (51) which reported low coverage to drug uptake to be associated with living in CDTI program area for less than five years. Ulanga district has vast fertile land and precious minerals deposits, such as spinel, gold, ruby and graphite that act as a natural factor for emigration that define its unique context and explains the existence of many community members who are not native. If these unique features are not well accommodated during MDA programs, it hampers the effective implementation and attainment of optimal MDA coverage.

The study found low treatment coverage below the WHO recommended coverage of 85% for successful transmission interruption (72). Poor treatment coverage in areas which have been in the MDA program for a long period have been reported earlier, by other research studies (17,48,62) as a probable barrier to transmission interruption. Understanding these dynamics and instituting corrective measures may form the basis for successful MDA programs and sustaining efforts to achieve high treatment coverage.

Drug uptake was observed to be high among participants who were influenced by possible prevention effects of Ivermectin towards Onchocerciasis (p<0.001). This could have been attributed to decreased number of people with visible signs of Onchocerciasis in the community thus increasing the community’s belief in the impact of the drugs. Studies from Cameroon and Bukinafaso reported that about three quarters of the people who complied to CTDI believed in the effectiveness and its health benefits of the Ivermectin drugs (8), (66). In the current study it was established that low drug uptake was associated with fear of Ivermectin side effects. This may be due to misconceptions and myths about IVM drugs that exist in the communities such as attributing Ivermectin effects to male impotence, birth control and swelling thus resulting in poor acceptance of preventive services (65). Knowledge of MDA distribution cycles was found to be an important determinant of drug uptake. This knowledge increased the possibility of participation in several distribution cycles compared to those who lacked the knowledge on distribution cycles. This situation could have been due to lack of effective awareness campaigns leading to inadequate community sensitization before drug distribution during the MDA cycle. The findings from this study on the positive contribution of community knowledge of MDA distribution cycles on drug uptake; are contrary to earlier research that reported limited association between drug uptake and knowledge of MDA rounds. Investing into intensive community directed health education and sensitization would result into increased drug uptake and paving the way towards elimination. (41). There is a possibility that these intrinsic characteristics such population mobility and livelihood activities for drug recipients within Ulanga district were overlooked, therefore explaining consistent low coverages reported by both program and various study surveys within the district. It is therefore important that these factors are well articulated in MDA campaign to attain recommended coverage and accelerate the pace towards control and elimination.

## Conclusion

This study has demonstrated low coverage of drug uptake as recorded in the community drug distribution register which indicates that the effectiveness of the MDA activities was not up to the recommended level. The longer the person lives in the community the higher the chance of compliance to drug uptake. Young adults contributed positively to drug uptake unlike elders. Belief in the preventive effects and understanding the pattern of MDA distribution interval contributed positively to drug uptake while fear of Ivermectin side effects affected individual participation and compliance to drug uptake. Misconceptions about ivermectin drugs specifically the belief that these drugs cause impotence and sterility, swelling as well as death affected participation in the MDA program and compliance to drug uptake.

### Recommendations

To improve future compliance to drugs uptake it is recommended that innovative approaches to social mobilization through community led health education campaigns and integration with existing health promotion should be considered not only to address drug misconceptions but also to intensify awareness of the benefits of ivermectin in Onchocerciasis control and MDA distribution cycles.

## Study Limitation

The main limitation of this study was the fact that investigations were conducted four to five months after the last MDA distribution paving a way for the recall bias. Nevertheless, it is believed that the results concerning the treatment coverage and compliance to drug uptake are reliable; because the methodological approach included review of all the records available in the community register of the study area and cross checking with the records at the district level. Also, the uniqueness of ivermectin tablets themselves in terms of both physical appearance (small and white) and their distribution strategy makes the treatment cycle easily remembered.

## Data Availability

We fully consent that data used in this study will be made available.

## Acknowledgement

This study received financial assistance from Tanzania Field Epidemiology and Laboratory Training Program and the Neglected tropical Diseases Control Program, Tanzania. We are immensely grateful to the Neglected Tropical Diseases Control Program for availing space in the Nadat base and equipment for electronic data collection.

